# The experiences and perceptions of working with Anaesthesia Associates: a survey of UK anaesthetists in training

**DOI:** 10.1101/2023.05.22.23290109

**Authors:** Ben Evans, Leyla M Turkoglu, James Brooks, Jeevakan Subramaniam, Stuart Edwardson, Roopa McCrossan, Naomi Freeman, Danny J. N. Wong

**Author notes:** **Corresponding Author:** Dr Ben Evans, Freeman Hospital, Freeman Road, NE7 7DN.

## Abstract

The future and sustainability of the anaesthetic workforce is a growing concern with a projected shortfall of 11,000 anaesthetists by 2040. The supply of anaesthetists able to provide safe anaesthesia care does not meet the rising demand of surgical waiting lists. In recent years, changes to recruitment and curricula for anaesthetic specialty training have resulted in significant bottlenecks to training progression, further compounding the deficit in the numbers of fully trained anaesthetists. A rapid expansion the Anaesthesia Associate (AA) workforce has been proposed as one of the solutions to this worsening gap. However, no robust analysis of the impact of the expansion in AA numbers on anaesthetists in training has been conducted. There remain a number of concerns regarding access to training experience, scope of supervision, out of hours workload, equity of pay and cost of training, as well as the impact on future numbers of anaesthetists in training. In order to help shape the future integration of this workforce, we surveyed current anaesthetists in training, asking about their experiences of working with AAs, and their perceptions of the future expansion and regulation of these associate professionals. Through an online survey, we collected both quantitative and qualitative data to give a thorough representation of anaesthetists in training experience. Our results confirm that the impact of AAs on training remains a polarising topic. A third of anaesthetists in training with prior experience of working with AAs reported a negative impact on their training experience. Factors mentioned included lack of case numbers, lack of access to learning and performing regional anaesthesia, and lack of clarity in lines of supervision. Of those with no prior experience working with AAs, there was a strong negative perception towards expansion of the workforce. A small proportion described a positive experience, indicating that with clear role definition, careful implementation along with co-operation, a positive experience in all departments could be achievable. Our findings suggest a need for increased consultation and communication with stakeholders in the anaesthesia workforce, including anaesthetists in training, to ensure smooth and safe integration of the AA workforce.

## Introduction

Anaesthesia is the largest hospital-based specialty in the United Kingdom. By 2033, it is predicted that there will be a shortfall of 4,000 consultant anaesthetists [1]. At this time, the UK is not training enough anaesthetists to fulfil the even further projected shortfall of 11,000 by 2040. The cessation of elective operations during the COVID-19 pandemic has led to a backlog of over 7.2 million patients waiting for consultant-led elective care [2], with the vast majority of these procedures requiring an element of care delivered by anaesthetists. The government plans to tackle these long waits in their plan for the NHS [3]. It has been suggested that increasing the numbers of Anaesthesia Associates (AAs) (known previously as ‘Physicians’ Assistant in Anaesthesia’, and originally ‘anaesthetic practitioners’ when the role was devised) could be used to help meet this significant clinical demand [4].

AAs were introduced in 2004, and are part of the Medical Allied Professionals group. They are non-medical anaesthetic practitioners who work under the supervision of a consultant anaesthetist [5]. AAs are currently unregulated, however there are plans for regulation by the General Medical Council (GMC) to commence in 2024. As part of their regulation, the GMC requires a generic curriculum, which the Royal College of Anaesthetists (RCoA) has contributed to via the Faculty of Anaesthesia Associates Founding Board (FAAFB) and the Anaesthesia Associates Curriculum Review Group [6]. In 2016, the Association of Anaesthetists and the RCoA co-produced a scope of practice document which outlined the clinical settings in which AAs could work, and the procedures they could perform [7]. This scope of practice document proposes direct supervision of AAs for induction and emergence of anaesthesia, as well as limiting their clinical practice to exclude acutely ill, obstetric, paediatric patients and regional anaesthesia. The Association of Anaesthesia Associates states the 2016 scope of practice is a starting point and AAs can work beyond this scope under a locally agreed governance structure [8]. This document is currently being revised to incorporate the expansion of the AA role.

The introduction of AAs was initially met with concerns regarding safety as well as the impact on anaesthetists in training [9,10]. This impact has not been thoroughly studied since their introduction [11].

The planned increase in the number of AAs [12], which includes fully funded training places, is an opportunity to shape the work of this professional cohort to add diversity and flexibility to the workforce, without duplicating the anaesthetist’s role. The lack of evidence regarding the impact of AAs on the training experiences of anaesthetists leaves uncertainty as to how this may affect the physician anaesthetists of the future [9]. We aim to increase this evidence base by asking UK anaesthetists in training about their experiences of working with AAs, and the effect of upscaling this professional cohort. We also surveyed anaesthetists in training who do not have experience working with AAs to ascertain their perceptions. This may give an idea of how this professional cohort could be smoothly and safely expanded into the current workforce landscape.

## Methods

Our survey was constructed using a modified Delphi approach [13]. An initial pilot questionnaire was collaboratively prepared by the authorship—a mixture of anaesthetists of varying career stages, including those in training. Feedback from this pilot survey was used to refine the questions to limit lead bias. The final survey was designed with embedded survey logic enabling streaming of respondents depending on whether they had prior experience working with AAs. Those who had previous or current working contact with AAs were asked specific questions about their experience, any impact on their training, and observations regarding AAs scope of practice. Both those with prior experience working with AAs and those without were then asked the same questions later in the survey regarding future changes to the AA workforce, supervision and regulation. These further questions were directed to all respondents—both those with and without previous experience working with AAs—regarding the potential impact on anaesthetists in training, if further development of the AA role beyond regulation were to proceed. Respondents were asked separate questions about the perceived impact of AAs practising independently with distant (out of hospital) consultant supervision, and working out of hours.

The survey was distributed as a Google Form (Mountain View, CA, USA) between 9^th^ January 2022 and 9^th^ February 2022 via social media platforms and the trainee networks of the Association of Anaesthetists and the RCoA. The Heads of School for Anaesthesia in each postgraduate training region were contacted and asked to distribute the survey amongst their body of anaesthetists in training. The survey is available in the online Supporting Information (**Appendix S1**).

To ensure authentic responses were collected, respondents were asked to provide their email addresses when responding. After manual inspection of these addresses, a follow-up email was sent. Any responses with email addresses that returned a non-delivery report/receipt (NDR), or which appeared obviously false, were removed in their entirety.

We report descriptive statistics of the respondent characteristics, and numbers and proportions of categorical responses to survey questions. Thematic analysis of the free text comments was performed by the authors using the method recommended by the Association for Medical Education in Europe [14]. Quantitative analysis was conducted in Microsoft Excel and R version 4.2.2 (R Foundation for Statistical Computing, Vienna, Austria). Sentiment analysis was performed using the SentimentR R package, with a method described by Rinker [15]. This sentiment analysis approach allows the calculation of text polarity by dictionary lookup of terms while also incorporating valence shifters.

## Results

A total of 663 survey responses were received. Seventeen responses were removed from analysis (15 due to an invalid email address, and 2 were not anaesthetists in training), leaving 644 responses for thematic analysis (**Figure 1**).

**Figure 1:**
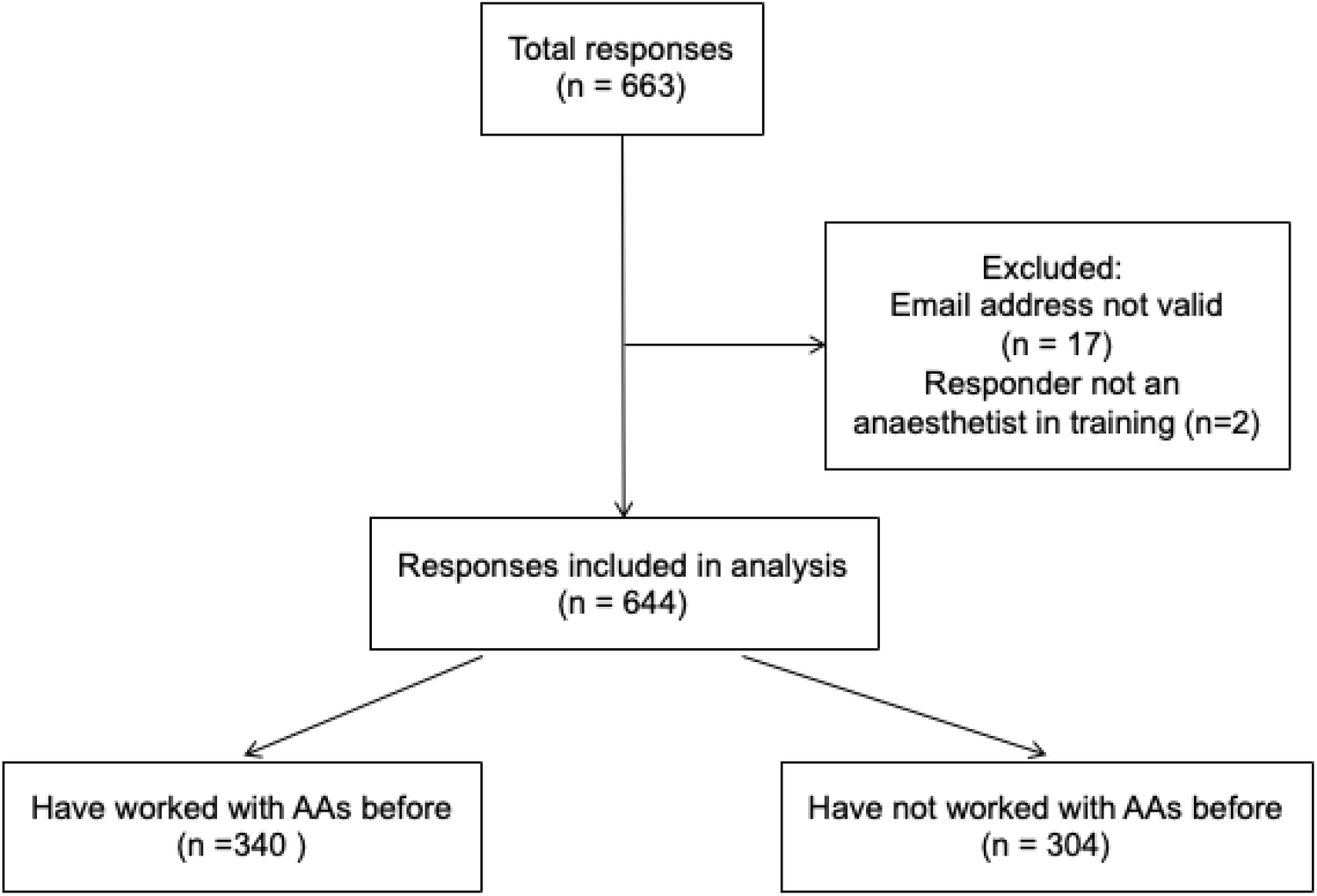
Flowchart of responses included in analysis.

Of the valid responses, 340 (52.8%) had worked with AAs before, while 304 (47.2%) did not have direct experience working with AAs.

The characteristics of respondents are shown in **Table 1**. There was a spread of respondents in terms of stage of training with 299 (46.4%) from Stage 1, 195 (30.3%) from Stage 2 and 149 (23.1%) from Stage 3. The distribution of responses from different training grades did not significantly differ between those who had previous experience working with AAs and those who did not. Responses were received from every deanery within the 4 devolved NHS nations.

**Table 1:**
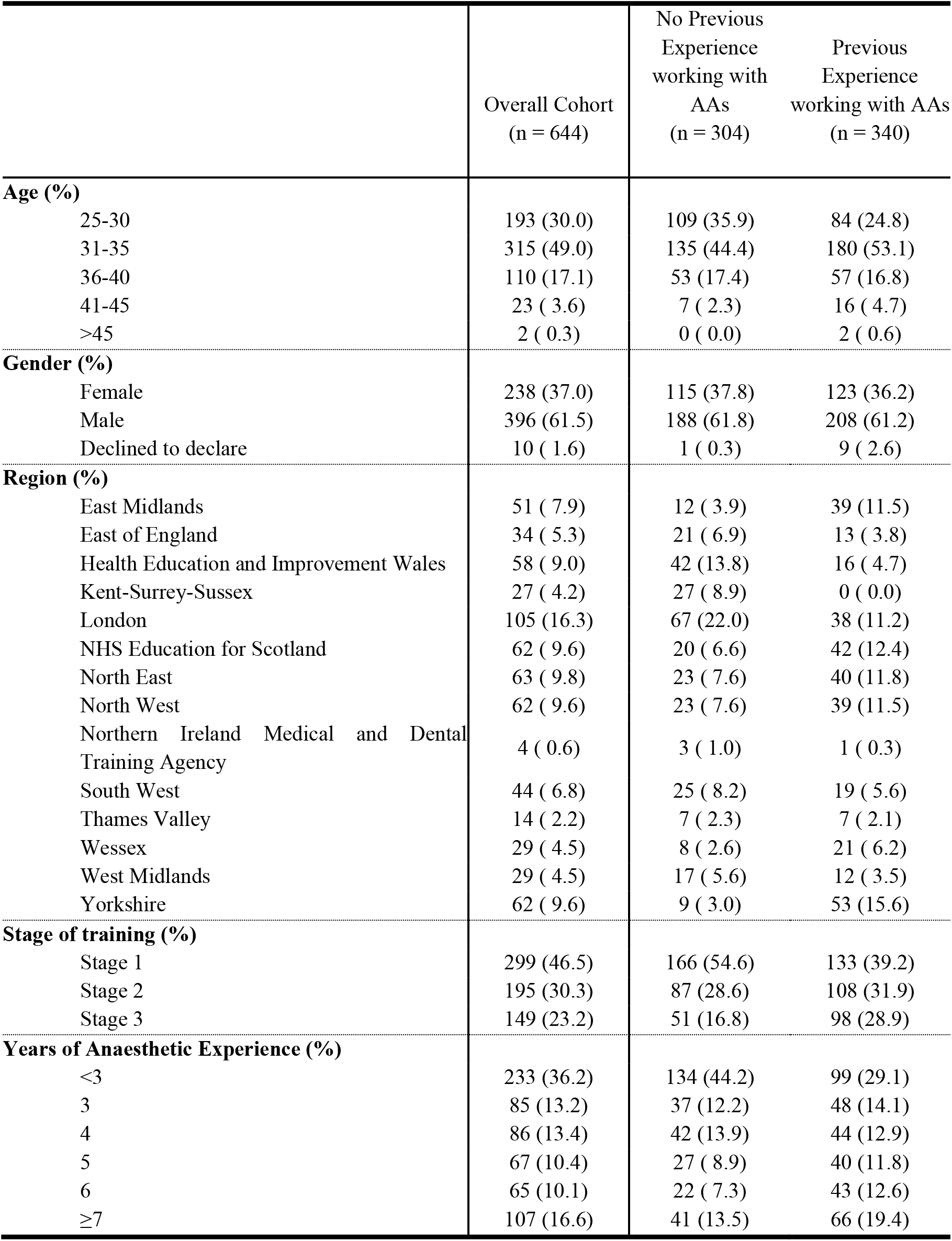
Characteristics of the 644 respondents included in analysis. Values are number (proportion).

### Anaesthetists in training who have worked with AAs

Anaesthetists who had previous experience of working with AAs (n = 340) were asked about the impact on their training. A higher proportion of respondents perceived the impact on their training to be negative than positive (**Figure 2**).

**Figure 2:**
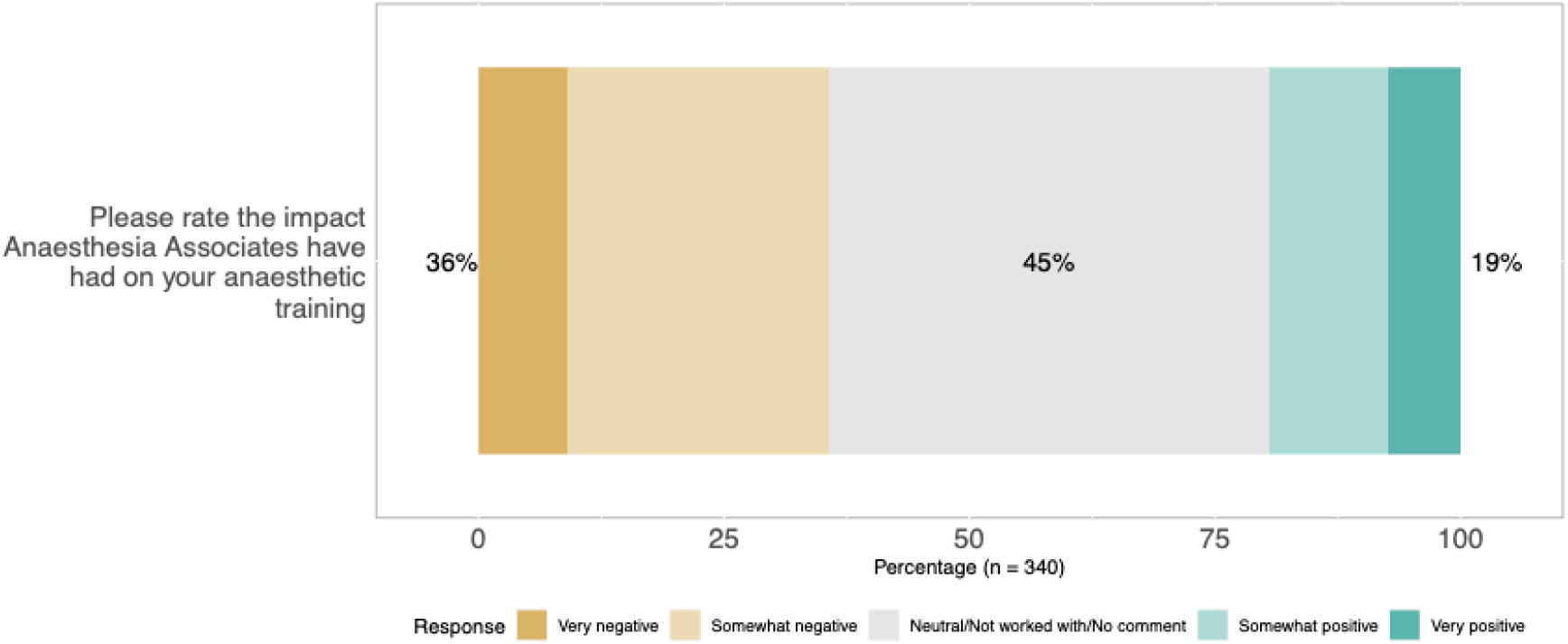
Responses from anaesthetists in training with previous experiences working with AAs to the question, “Please rate the impact Anaesthesia Associates have had on your anaesthetic training.” A higher proportion of anaesthetists in training reported that AAs impacts on training were either ‘very negative’ or ‘somewhat negative’ (121/340 = 35.5%), compared to those who reported ‘very positive’ or ‘somewhat positive’ impacts (66/340 = 19.4%). Approximately 45% said the impact was neutral or had no comment.

The same anaesthetists were asked whether they had witnessed AAs working beyond the 2016 joint scope of practice guidance. One hundred and forty-four (42.3%) respondents were not sure, 64 (18.8%) stated they had seen AAs work beyond this guidance, and 131 (38.5%) stated they had not.

Further details about stated divergence from this scope of practice was then clarified via additional follow-up questions. Two hundred and sixteen (63.5%) free-text responses regarding divergence from scope of practice were received and categorised into common themes. The most common response was AAs ‘performing regional blocks’ and then that AAs were not supervised in line with the guidance either preoperatively, intraoperatively or on emergence. The remaining respondents chose not to answer this question.

### Anaesthetists in training - all respondents

The majority of anaesthetists in training felt the impact of working with distant supervision would be negative, and responses were consistent regardless of whether respondents had prior experience working with AAs (**Figure 3**). Similarly, a majority of respondents perceived the impact of AAs working out of hours to be negative, however, the responses to this question showed variation between those with and without prior experience working with AAs—32.3% of trainees with experience working with AAs viewed this positively, compared to 20.7% of those with no prior experience working with AAs.

**Figure 3:**
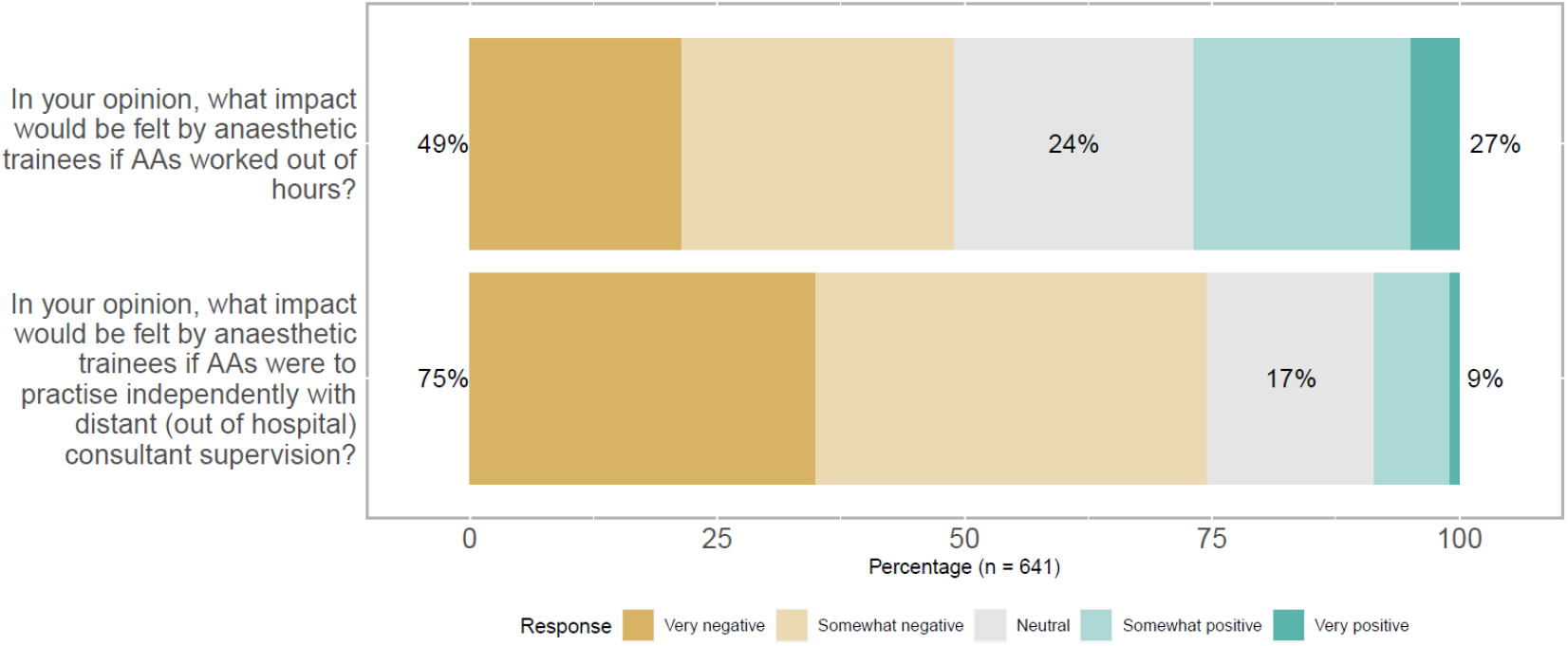
Responses from all anaesthetists in training, regardless of whether they had previous experience working with AAs, to questions about the perceived impact on their training if AAs were to undertake out of hours work or work under distant supervision. We received responses from n = 641 respondents to both these questions, with 3 declining to respond. Forty-nine percent of respondents felt the impact of AAs on anaesthetic trainees would be either ‘very negative’ or ‘somewhat negative’ if AAs worked out of hours, vs. 27% who responded either ‘very positive’ or ‘somewhat positive’. Similarly, the majority (75%) felt the impact on trainees if AAs were to practice independently with distant consultant supervision would be either ‘very negative’ or ‘somewhat negative’, compared with 9% who perceived this to be either ‘very’ or ‘somewhat’ positive.

Six hundred and forty-one individuals responded when asked about the prescribing capabilities of AAs. The most commonly selected answer was ‘limited formulary, e.g. anaesthetic and peri-operative medicines only’ 47.1% (302). 30.7% (197) thought AAs should have ‘no prescribing capability’, while only 16.6% (107) felt that AAs should have full prescribing rights. These responses were consistent across the different grades of anaesthetists surveyed.

Respondents were then asked to rate the priority of training for different groups given the current backlog of elective surgery. They were asked, using a Likert scale, to rate the importance of training ‘Higher Anaesthetic Trainees’, ‘SAS doctors’ and ‘Anaesthesia Associates’. 71.5% (461) felt that training Higher (Stage 3) anaesthetic trainees was ‘very important’, with 2% (14) deeming it ‘fairly important’ (the second highest descriptor on the Likert scale) and 25.0% (161) ‘important’. Only 1.6% (8) deemed it ‘slightly important’ or ‘not important at all’ (the two lowest descriptors). Expansion of SAS doctors was similarly deemed ‘very important’ by 54.8% (253), with 24.6% (159) deeming it ‘fairly important’ and 24.5% (158) ‘important’. 11.8% (72) deemed the expansion of SAS doctors ‘slightly important’ or ‘not important at all’.

Support for an expansion of AAs in the anaesthetic workforce was assessed using the same method. All respondents answered this question. 36.0% (232) were ‘Somewhat against’ expansion of the AA workforce, while 30.4 (196) were ‘Totally against’. 18.0% (116) remained neutral, while 12.1% (78) were ‘Somewhat for’ and 3.6% (23) ‘Totally for’. When splitting these results by exposure to AAs, 60.8% (207) of those who had worked with AAs were broadly against (combined ‘Totally against’ and ‘Somewhat against’) versus 72.4% (220) of those without experience with AAs.

## Thematic Analysis

Thematic analysis of responses from anaesthetists in training who had previously worked with AAs were assessed; there were 207 responses with 62.8% (130) negative, 19.3% (40) positive and 17.9% (37) neutral. The most common negative themes were ‘Poor or Unclear Supervision’, 12.6% (26), ‘Loss of Regional Anaesthesia Experience 10.1% (21) and ‘Trainee covering emergency work so AAs can do elective work 7.7% (16). The most common positive themes were: ‘AA’s delivering teaching’ 8.7% (18), and ‘AAs freeing up consultants to teach trainees’ 2.9% (6).

Of those without previous experience working with AAs, there were 103 responses with 96 (93.2%) negative, 4 (3.9%) positive and 3 (2.9%) neutral. The most common negative themes were ‘Lack of Training Numbers’ (n=49, 47.6%), ‘Reduced Training Experience’ (n=21, 20.4%) and ‘Supervision and Safety Concerns’ (n=13, 12.6%). There were too few positive comments to coherently categorise.

Indicative quotes from free text analyses are presented in **Table 2** exemplifying positive and negative perceptions relating to the direct impact of AAs on training opportunities and experience uncovered in thematic analysis. **Table 3** shows quotations from analysis of the thematic analysis related to broader negative perceptions, there were no positive perceptions regarding these topics uncovered in analysis.

**Table 2:** Example quotes relating to the direct impact of AAs on training opportunities.

| Positive Perceptions |  | Negative Perceptions |  |
| --- | --- | --- | --- |
| Prior experience working with AAs | No prior experience working with AAs | Prior experience working with AAs | No prior experience working with AAs |
| ‘Great learning resource, especially as a novice trainee.’<br>*Stage 1, Scotland.*<br>‘Very helpful and good at teaching and often do lots of the elective orthopaedic lists to free up teaching opportunities for trainees.’<br>*Stage 1, North East England.*<br>‘I found it helpful as it meant the consultant could focus more on teaching me as the AA was looking after the patient.’<br>*Stage 3, London.*<br>‘At no point have I missed out on a learning opportunity because of an AA, and this includes when I worked in a hospital with trainee AAs during the height of the Covid pandemic when theatre activity was significantly reduced.’<br>*Stage 3, Scotland.* | ‘I think the limited and supervised covering of simple lists by AA’s is a good solution to providing service delivery.’<br>*Stage 2, Kent-Surrey-Sussex.*<br>‘AAs definitely will be an additional help, for quicker turnover of cases, as an extra pair of hands, but have to bear in mind that anaesthetic training cannot be fully comparable to AA training.’<br>*Stage 2, Yorkshire.* | ‘They require training but we also need training. We need to know list management and I have felt, in the past, qualified AAs get lists and we get micromanaged.’<br>*Stage 3, Scotland.*<br>‘Departments, especially consultants, sometimes prioritise teaching and training them as they become permanent members of staff and trainees move on.’<br>*Stage 1, Scotland.*<br>‘They were very friendly and welcoming although definitely took training opportunities away from me as a trainee.’<br>*Stage 2, Yorkshire.* | ‘Would fear the training opportunities lost from trainees and the departmental shift to train those who will not be rotating every 6 months but are permanent members of the team. Would fear lack of exposure to as much variety’<br>*Stage 1, North East England.*<br>‘I worry PAs would be trained up to do regional work, lines etc and yet more opportunities taken away from trainees, or do "simple" ASA 1/2 daycase lists, and yet more training opportunities for CTs lost.’<br>*Stage 1, Scotland.*<br>‘Core trainees struggling to fulfil basic competencies for IAC due to lack of cases.’<br>*Stage, 1, Scotland.* |

**Table 3:**
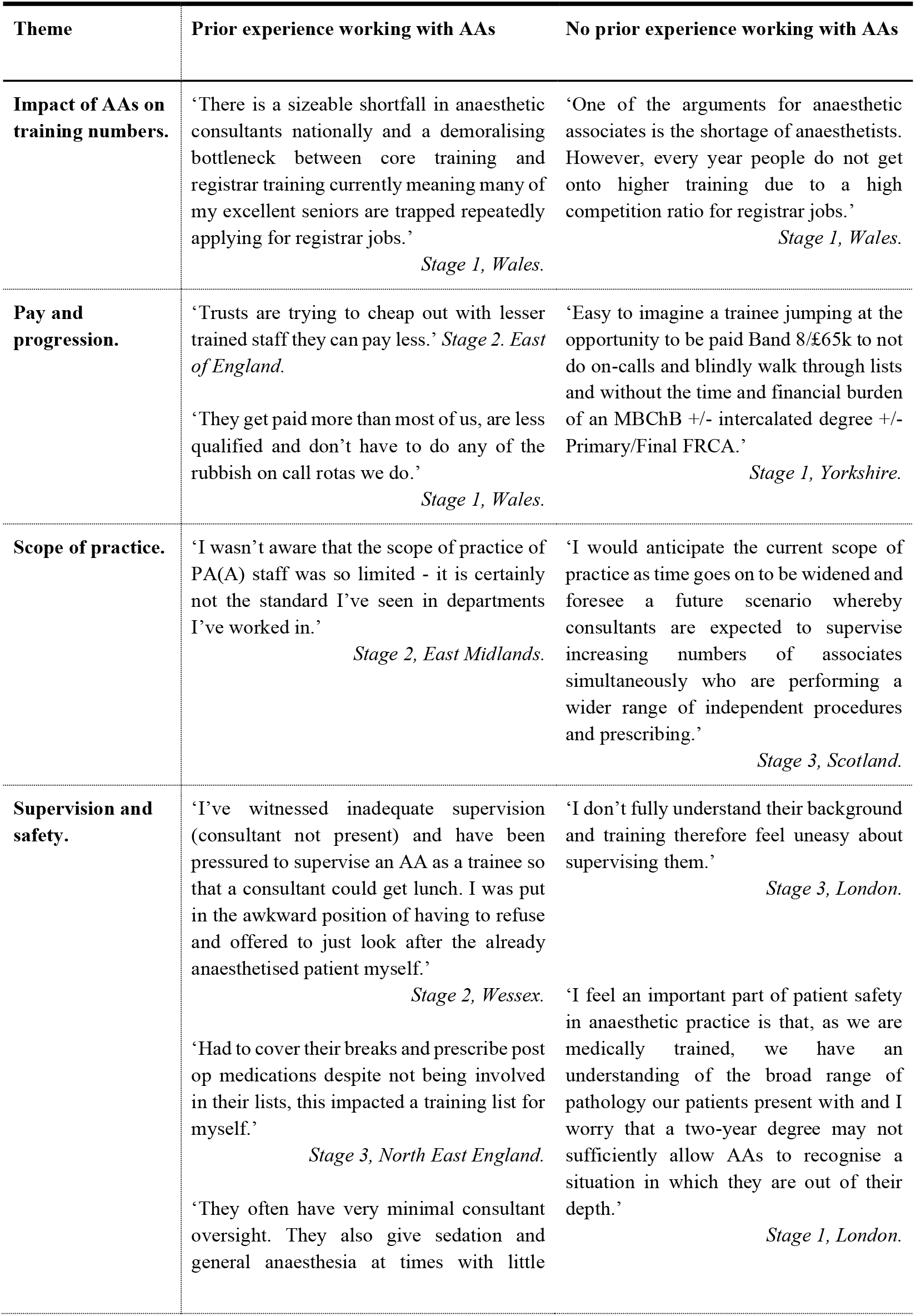
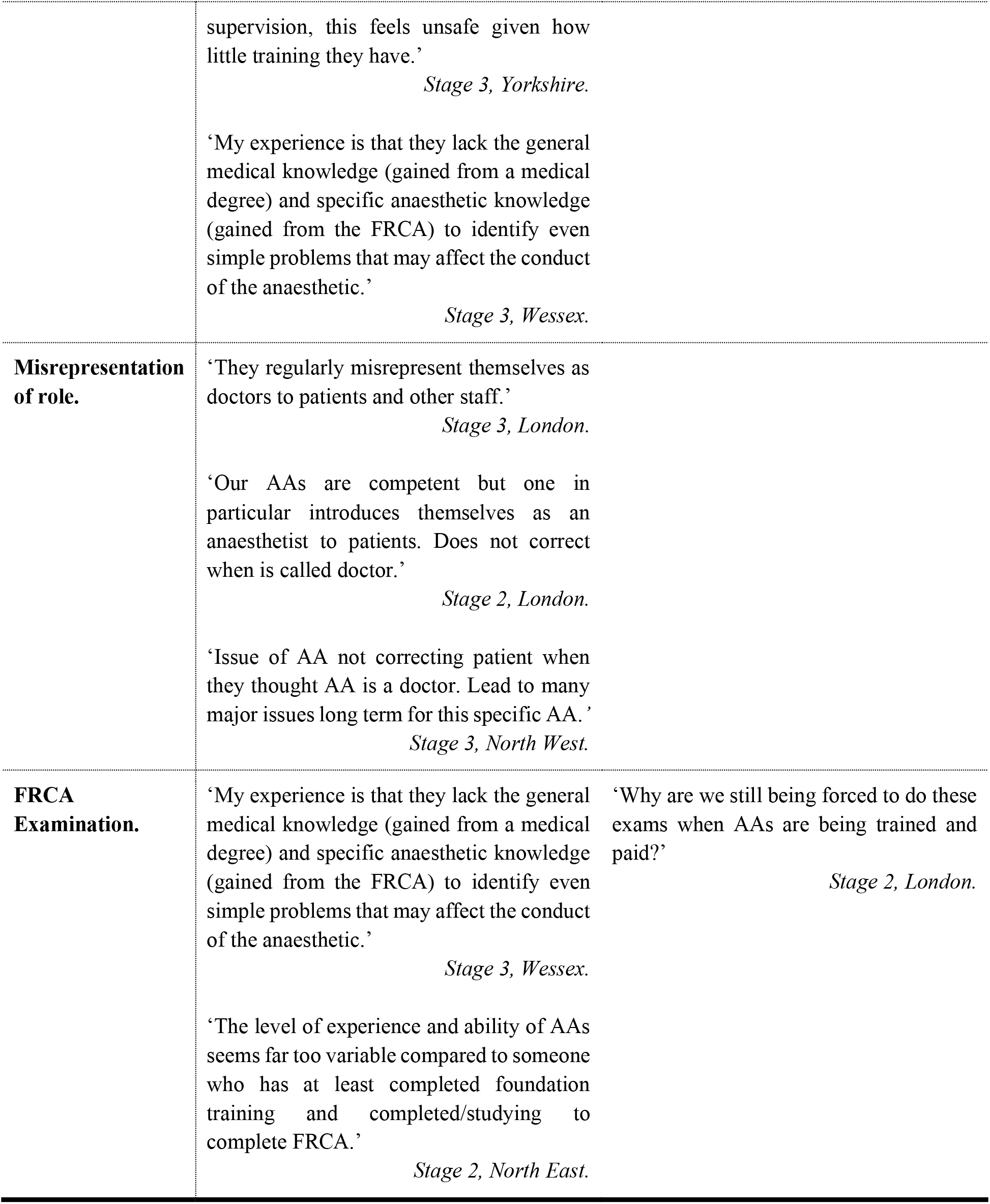
Negative perceptions relating to themes uncovered in thematic analysis of free text comments. There were no positive perceptions uncovered relating to these themes.

The indicative quotes in **Table 3** highlight the common issues that anaesthetists in training have experienced or are concerned about; themes included the pay of AAs being over and above many anaesthetists in training when on-call supplements were removed, the preferable working patterns of AAs compared to anaesthetists in training, a lack of clarity about supervision, and confusion regarding AAs being mispresented to patients as physician.

Some respondents referenced workforce deficits of anaesthetic assistants (operating department practitioners and anaesthetic nurses) and a concern that an expansion of AAs would further detract from this skilled workforce and thus leave anaesthetic departments struggling for safe staffing levels.

The Fellowship examination of the RCoA (FRCA) was raised by many respondents, with comments indicating how AAs were able to practise without the FRCA, whilst anaesthetists in training were required to obtain the FRCA to complete their training. Although anaesthesia associates will have to pass the AA Registration Assessment (AARA) for full registration with the GMC, many felt aggrieved that they had to undergo rigorous post-graduate qualifications with learning in their own time while AAs did not.

Other respondents felt that given the increasing patient complexity, training AAs who would only be managing ASA I-II patients without direct consultant supervision [16], was not a good use of resources.

Sentiment analysis performed on free text responses to the survey from all respondents showed that attitudes towards AAs were generally positive (greater than zero, p<0.001) among respondents with previous experience working with AAs, with median (IQR [range]) scores of 0.094 (−0.002–0.237 [-0.750–1.366]) in this group. The median (IQR [range]) sentiment scores of respondents with no previous experience working with AAs was 0.014 (− 0.079–0.123 [-0.625–0.794]), and not significantly different (p=0.230) to zero (neutral). The difference in sentiment scores between both groups was statistically significant (p<0.001), and persisted after adjustments were made for age, gender, stage of training and region of the respondents in a multivariable linear regression model. Variance in sentiment scores was high in the responses from both groups, suggesting wide variation in opinion (**Figure 4**).

**Figure 4:**
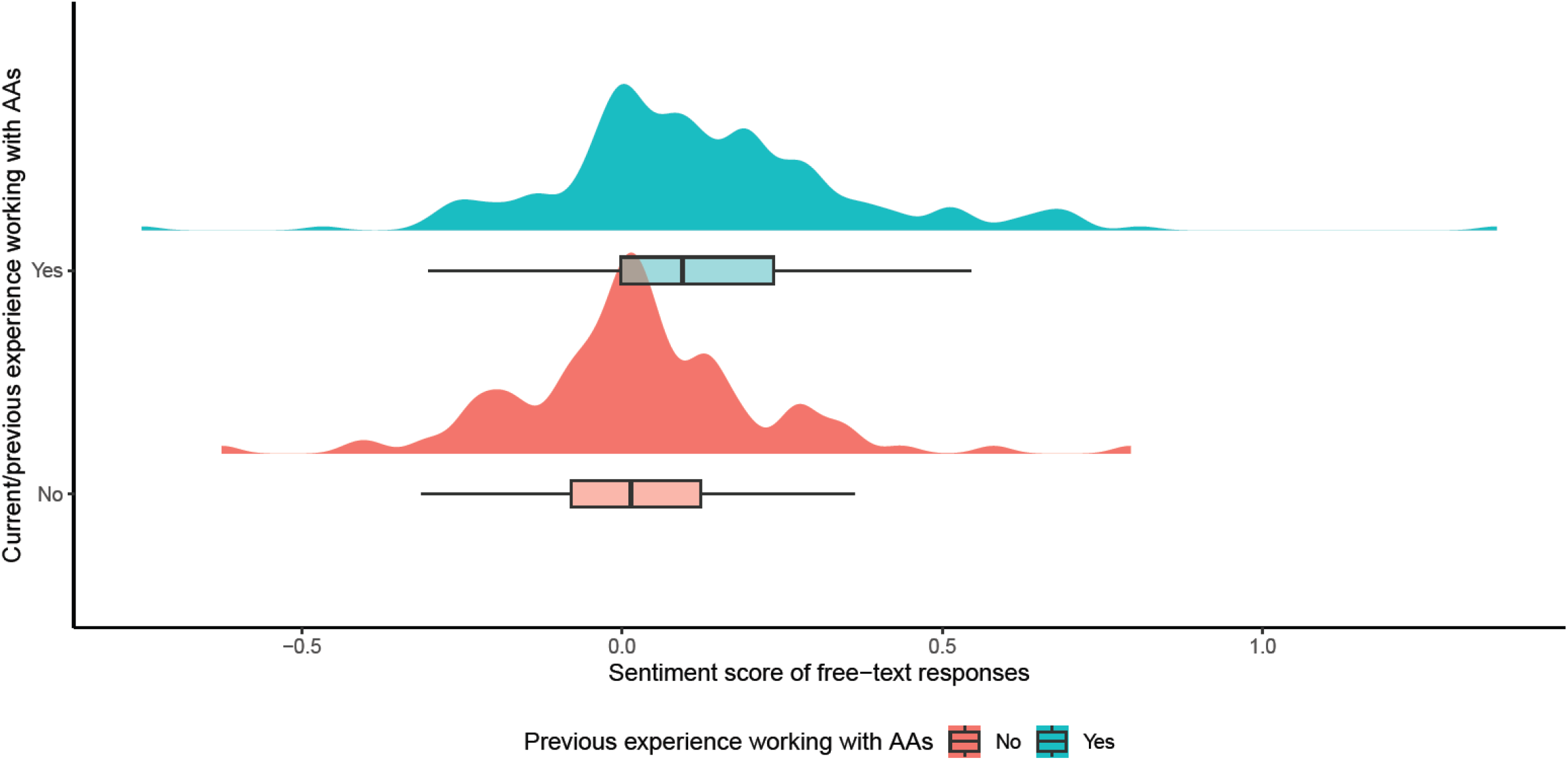
Distribution of sentiment scores calculated from free text responses. These sentiment scores were stratified by whether the respondent providing free-text responses had prior experience of working AAs, or no prior experience. The sentiment scores were higher in responses from those who had prior experience working with AAs than those who did not (p <0.001).

## Discussion

In this survey, we report the views from a large sample of anaesthetic trainees on AAs and their impact on anaesthetic specialty training. While many respondents were neutral on the impact of AAs on training, around a third of respondents with previous experience working with AAs felt that they had negatively impacted training. Anaesthetists in training were more likely to perceive the impact of AAs negatively than positively, when asked about AAs practising independently with distant supervision, or working out of hours. A higher proportion of respondents were in Stage 1 training compared to those in Stage 2 and 3 [1]. Thematic analysis of free-text responses revealed negative themes around supervision of AAs and competition for certain training opportunities; alongside positive themes such as additional resourcing to free up anaesthetists to perform tasks and provide training. A majority (irrespective of prior work with AAs) were against the expansion of AAs in the workforce in the near future. A minority of those with experience working with AAs held this view. Sentiment analysis demonstrated a small positive sentiment overall in the free-text survey responses, with a significantly more positive sentiment score among respondents with previous experience working with AAs.

There are 4,311 Anaesthetists in Training, our survey had 644 responses (14.9%) [1]. This is considerably more than previous studies but there remains a large proportion of trainees whose views are unknown. This cohort of anaesthetic trainees has been thoroughly ‘surveyed’ over recent years due to the recent Covid-19 pandemic as a way of assessing effects of recruitment, curriculum and working changes and thus ‘survey fatigue’ may be implicated [17]. However, the sample of respondents is representative of the entire UK training cohort, when compared to the RCoA medical workforce census report [1], we show both an even geographic spread across the UK training deaneries and the proportion of respondents at each training stage reflects the national training picture [1]. Three hundred and forty respondents worked with AAs before. There are currently 173 AAs registered with RCoA and working within the UK [1]. This respondent number therefore reflects a significant proportion of trainees who have worked with this relatively small group of AAs.

The impact of AAs on training is polarising. There are negative perceptions that must be addressed before widespread deployment of AAs should be supported. In particular; access to training opportunities, regulation and supervision. The new regulation and scope of practice documents for AAs should seek to clarify these perceptions alongside expansion of the workforce. Auspiciously, those who have worked with AAs before have a less negative perception than those who have not. For some anaesthetists in training, AAs are positively perceived and their impacts on training have been beneficial. This improvement in the training experience for physician anaesthetists should be welcomed but is not the case across the UK.

This survey showed clear demand for an increase in anaesthetic training numbers, as well as support for developing SAS careers. This is driven by significant workforce pressures and training bottlenecks. The recent, and now regular, lack of specialty training numbers thereby potentially cutting short the early careers of those already partially trained in anaesthesia further compounds this [18]. There is certainly no limitation in enthusiasm for doctors to become anaesthetists, given that the competition ratio to enter training was 4.2 in 2022 [19]. The results of this survey strongly suggest that the current anaesthetic trainee workforce do not think that expansion to the AA workforce alone will improve workforce issues. Some free-text comments point to this as a method of clearing surgical backlogs for reduced cost. This seems to go directly against the planning guidance for the introduction and training for AAs which states *“It is not about … cost cutting, or simply to address staff shortages; it is ensuring that the service user receives the most appropriate care, at the most appropriate time from the most appropriate person”* [16].

Aside from their experience of training, many respondents described actively comparing their pay and working conditions (including out-of-hours working patterns) with AAs. There was a strong sense of dissatisfaction with this, citing the number of years of training medical anaesthetists require, while also having to work unsociable hours. Given the current national industrial action from junior doctors this needs to be considered in context, and may well represent general discontent as opposed to an opinion regarding AAs [20].

Within current guidance, the AA workforce are only able to take part in anaesthetic management of patients who are ASA I-II undergoing minor to intermediate surgery only. However, a recent large-scale contemporary summary of the characteristics of the surgical population suggest that this patient cohort is decreasing. Since 2013, mean age, BMI, and comorbidity burden of all patients has increased. The proportion of ASA I patients has decreased from 37% to 24%. This equates to an all-cause mortality increase of 27% [21]. This increasingly complex and higher risk perioperative workload is highly likely to be reflected within those patients who have spent significant time on operative waiting lists.

Anaesthetists in training were broadly against the expansion of AAs to work out of hours and with indirect (distant) supervision. This is perhaps due to a significant perceived overlap, were this to happen, between the role of AAs and anaesthetists in training and concerns regarding exposure to clinical opportunities. This sentiment was also commonly intertwined with concern regarding confusion around supervision in the case that both medical anaesthetist and AA are present in hospital with consultant supervising from home. In addition to concerns regarding clarity of supervision, our respondents raised concerns regarding lack of regional anaesthesia experience if working in a department with AAs. This is of particular concern given the new emphasis on independence with a broadened range of regional anaesthetic techniques now stipulated in the new RCoA curriculum [22].

There is little previous research on this topic, and a search of the literature revealed only 2 studies. A 2015 trainee survey conducted by the Group of Anaesthetists in Training of the Association of Anaesthetists found that 82% of the 399 trainee respondents were concerned by the perceived negative impact of AAs on postgraduate specialty training [23]. A separate qualitative study which interviewed 7 anaesthetists in training in one health board found largely positive feedback [9]. A number of other peer reviewed articles make reference to the impact on anaesthetists in training from AAs. The RCoA bulletin, in 2014, describes the impact of PA(A)s at one NHS trust citing “*many examples of an actual benefit to junior doctors in training”* [24]. An Association of Anaesthetists (then AAGBI) report on Physicians’ Associates in Anaesthesia, the only comment regarding impact on Anaesthetic trainees was “*The majority felt that they impacted positively, allowing their trainers more freedom to train*.*”* [11] Conversely, an article from 2007, around the inception of anaesthetic practitioners, discussed the potential for an adverse effect on medical anaesthetists and concluded, “*APs will be able to do some of the work left by the reducing number of medical anaesthetic trainees”* [25]. By comparison, this survey is the largest and most broad-ranging collection of evidence on this topic. Importantly, we targeted respondents representing the entirety of the UK training deaneries irrespective of their experience of AAs. While the results from those with experience gives us a thorough idea of their views, those without experience have given us a strong idea of what perceptions there are of expansion of the AA workforce. This not only gives opportunity to shape upcoming regulation, but also framing future maintenance of training quality and advocacy for training numbers in the knowledge of these perceptions and hesitations.

With the upcoming regulation of AAs, it is imperative that the views of Anaesthetists in Training are taken into account on both a local and national level. Locally, College Tutors should be responsible for ensuring that training opportunities are not lost to departments that start to train AAs or deploy AAs to certain lists. Departments should be encouraged to undertake local quality improvement processes to ascertain this impact. Nationally, we suggest that the anaesthetic component of the GMC National Training Survey, should incorporate questions regarding the impact (both positive and negative) of AAs on training experience [26]. This could also be replicated in any future workforce census reports conducted by the RCoA. Ensuring the quality of training for anaesthetists is an active process, and cannot be assumed to be adequate in the face of a changing workforce and healthcare landscape. It is imperative that these questions are asked regularly, the results published, and with accountability.

While the expansion of the AA workforce has been led with reference to limited evidence of their positive impact on the training experience, our survey demonstrates that this has not been the case in a majority of circumstances. The reasons for this are likely to be complex and multifactorial. However, the minority evidence of positive impact does provide opportunity to learn from examples of good quality practice and regulation. Situations where this occurs should be identified and sought to be replicated nationally.

The anticipated national regulation guidance is sorely needed. The current 2016 Scope of Practice can be superseded in almost all departments in the UK. Our survey results suggest that this is indeed the case in many circumstances. Significant regional variation in expectations in training and practice will only worsen the potential for misunderstanding. Many of our free text comments described instances of AAs introducing themselves to patients as the ‘anaesthetist’. Given that at least 40% of the UK public do not know anaesthetists are doctors [27], the much-needed clarity of this regulatory document is an excellent opportunity to emphasise the background and skill-set of a medically trained anaesthetist. It is for this reason that we suggest that regulation includes that AAs be titled as Anaesthesia Associates, and specifically not anaesthetists.

Ongoing clear channels of communication between anaesthetists in training and anaesthesia associates should be constructed and maintained to ensure thorough understanding of each other’s roles and framework for practice.

## Conclusions

The future of the medical workforce is currently under intense scrutiny, and anaesthesia is no exception. Increasing the numbers of medical associate professionals will inevitably impact on the existing workforce, and some of these effects are demonstrated by our data. It is clear from the results of our survey that there are polarised attitudes towards Anaesthesia Associates. One third of those with previous experience working with AAs felt that they negatively impacted training. Anaesthetists in training were more likely to perceive the impact of AAs negatively than positively, when asked about AAs practising independently with distant supervision, or working out of hours. With increasing numbers of doctors in training leaving the UK or the profession entirely [28], it is vital that trainee views are taken into account at both a local and national level. Integration of the AA role in a pre-existing training model has not been without issue, and the real concern of loss of training opportunities for doctors is demonstrated in the survey responses. College Tutors have a role to ensure training opportunities for anaesthetists are not lost in favour of AAs. The scope of AA practice must be transparent for both anaesthetists and patients alike. It must be made clear that AAs are not doctors, nor anaesthetists, to avoid misplaced expectations. Supervision and scope of practice must be defined and adhered to.

Only by including the existing workforce in decisions about its future can positive change be made. The perspectives of the anaesthetists we surveyed show that, in the main, this has not happened and these changes have been imposed without proper forethought of the effect on training or trainer workload.

More work must be done to ensure a cohesive anaesthetic workforce can exist to deliver excellent, safe and reliable patient care.

## Supporting information

Appendix S1 - Questionnaire

## Data Availability

All data produced in the present study are available upon reasonable request to the authors

