## Appendix S1 - Questionnaire for "The experiences and perceptions of working with Anaesthesia Associates: a survey of UK anaesthetists in training"

### Survey of Trainee Experiences and Attitudes Toward Anaesthesia Associates in the UK

For all doctors currently in a UK anaesthetic training programme, we are keen to hear your views whether you have or have not directly worked with Anaesthesia Associates (AAs). We understand that many anaesthetists might consider themselves as trainees whether or not they are on a formal training programme, e.g. Trust-employed CT3 top-up fellows, we welcome your input in this survey as well.

Formal professional regulation for Anaesthesia Associates is currently being formulated by the General Medical Council. Numbers are expected to increase over the next three years. This has raised a number of important questions with regard to the potential impact on anaesthetic training.

AAs are healthcare professionals skilled in the administration and monitoring of many forms of anaesthesia. For more information, feel free to visit the following:
https://anaesthesiaassociates.org/general-info/faqs/
https://rcoa.ac.uk/training-careers/working-anaesthesia/anaesthesia-associates

AAs have been in clinical practice in the UK for many years. A number of departments anecdotally report positive experiences with regard to management of clinical workload and the effect on anaesthetic training. To clear the waiting list backlog, the current workforce situation must be improved. However, given reductions in elective operating and the significant lack of training numbers for doctors wanting to enter or continue in anaesthesia, there is no clear evidence on the impact of an increase in AA practice on anaesthetic training.

There is currently one UK paper, published June 2022 (available here: https://onlinelibrary.wiley.com/doi/epdf/10.1002/hpm.3502) which interviewed 7 trainees (6 of whom had worked directly with AAs) and found the impact to be generally positive. To this end, there remains no robust evidence base that centres the trainee voice.

We are a group of UK anaesthetists who are keen to gain a clear idea of trainee views or experiences of working with AAs in order that this feedback can influence the direction of AA regulation and integration into the workforce.

Thank you,

Dr Ben Evans
Dr Leyla Turkoglu
Dr James Brooks
Dr Stuart Edwardson
Dr Naomi Freeman
Dr Roopa McCrossan
Dr Jeevakan Subramaniam
Dr Danny Wong

* Indicates required question

1. If you consent to taking part and to having your responses analysed for research, publication and presentation, please provide your email address. By doing so you assert that you are an anaesthetist in training.

   Results from our survey will be reported in aggregate, and any responses will not be linked to your identity. We may contact you in the future to share the results of our survey with you directly or to ask you to participate in follow-up research.

   ____________________

2. What is your age?

   *Mark only one oval.*

   - <25
   - 25-30
   - 31-35
   - 36-40
   - 41-45
   - >45

3. How would you describe your gender?

   *Mark only one oval.*

   - Male
   - Female
   - Non-binary
   - Rather not say

4. In which region do you currently work?

*Mark only one oval.*

- East Midlands
- East of England
- Kent-Surrey-Sussex
- London
- North East
- North West
- South West
- Thames Valley
- Wessex
- West Midlands
- Yorkshire
- NHS Education for Scotland
- Northern Ireland Medical and Dental Training Agency
- Health Education and Improvement Wales

5. What is your current stage of anaesthetic training?

*Mark only one oval.*

- Stage 1
- Stage 2
- Stage 3
- Other: ____________________

6. How many years of anaesthetic training have you completed?

*Mark only one oval.*

- <1
- 1
- 2
- 3
- 4
- 5
- 6
- 7
- 8
- 9
- 10 or more

7. Do you currently work in, or have you ever worked in, a department with Anaesthesia Associates?

*Mark only one oval.*

- Yes
- No *Skip to question 12*

#### Questions about working with Anaesthesia Associates

8. Please rate the impact Anaesthesia Associates have had on your anaesthetic training

*Mark only one oval.*

- Very positive
- Somewhat positive
- Neutral
- Somewhat negative
- Very negative
- Not worked with/no comment

9. In your experience, have AAs worked to the 2016 scope of practice document published by the RCoA and Association of Anaesthetists? https://www.rcoa.ac.uk/sites/default/files/documents/2019-08/Scope-of-Practice-PAA-2016.pdf

*Mark only one oval.*

- Yes
- No
- Not sure

10. If AAs have worked beyond the scope of practice in the 2016 document (above), what wha ways has their scope differed? (tick all that apply)

*Tick all that apply.*

- [ ] Not supervised in line with scope of practice preoperatively
- [ ] Not supervised in line with scope of practice intraoperatively
- [ ] Not supervised in line with scope of practice on emergence/recovery
- [ ] Inadequate support staff (ie trained assistance)
- [ ] AAs prescribing medication
- [ ] AAs undertaking regional blocks
- [ ] AAs undertaking neuraxial blocks
- [ ] AAs undertaking paediatric practice
- [ ] AAs undertaking obstetric practice
- [ ] AAs undertaking initial airway assessment, acutely unwell or injured patient assessments
- [ ] Other: ____________________

11. If you have had experiences of working with Anaesthesia Associates as an anaesthetist in training, please tell us more below.

____________________

____________________

____________________

____________________

____________________

Questions about scope of practice and future planning

12. In your opinion, what impact would be felt by anaesthetic trainees if AAs were to practice independently with distant (out of hospital) consultant supervision?

*Mark only one oval.*

- Very negative
- Somewhat negative
- Neutral
- Somewhat positive
- Very positive

13. In your opinion, what impact would be felt by anaesthetic trainees if AAs worked out of hours?

*Mark only one oval.*

- Very negative
- Somewhat negative
- Neutral
- Somewhat positive
- Very positive

14. What prescribing capabilities do you think AAs should have?

*Mark only one oval.*

- Limited formulary eg anaesthetic and peri-operative medicines only
- All routine medications including patient's regular medications
- No prescribing capability or direct supervision of prescribing only
- Other: ____________________

15. Which of the below, in your opinion, are benefits of working with Anaesthesia Associates? (tick all that apply)

*Tick all that apply.*

- [ ] Reduction in non-theatre service provision (eg peripheral venous access service)
- [ ] Help train junior anaesthetic trainees
- [ ] Allow anaesthetic trainees to have more consultant contact for teaching
- [ ] Reduction of on-call commitment and burden for anaesthetic trainee rotas
- [ ] Allow a better balance of clinical exposure of both straightforward elective and complex emergency work for anaesthetic trainees
- [ ] Free up medical members of the department (including trainees) to perform other tasks suc as quality improvement, research, medical education etc
- [ ] Other: ____________________

16. Which of the below, in your opinion, are the drawbacks of working with Anaesthesia Associates? (tick all that apply)

*Tick all that apply.*

- [ ] Reduced airway management exposure
- [ ] Reduced theatre list management opportunities
- [ ] Reduced opportunity to perform regional anaesthesia procedures
- [ ] Reduced IAC training opportunities
- [ ] Reduced variation in speciality experience (including increased time in obstetrics and/or critical care
- [ ] Other: ____________________

17. How supportive would you be of an expansion of AAs in the anaesthetic workforce in the near future?

*Mark only one oval.*

- Totally against
- Somewhat against
- Neither for nor against
- Somewhat for
- Totally for

18. Given the backlog of elective surgery in the UK and the requirement for more anaesthetist please rate the priority of training the following groups to increase the workforce.

*Mark only one oval per row.*

| | Not important at all | Slightly important | Important | Fairly important | Very importar |
|---|---|---|---|---|---|
| **Higher Anaesthetic Trainees (stage 2/st4 to CCT)** | | | | | |
| **SAS doctors** | | | | | |
| **Anaesthesia Associates** | | | | | |

19. If you have not had experience of working with Anaesthesia Associates as an anaesthetis training, but have views not covered by the previous questions, please tell us more below.
